# Longitudinal Transitions in Initiation, Cessation, and Relapse of Smoking and E-Cigarette Use Among US Youth and Adults

**DOI:** 10.1101/2021.08.01.21261431

**Authors:** Eli Schwamm, Farzad Noubary, Nancy A. Rigotti, Krishna P. Reddy

## Abstract

**Introduction:** Longitudinal surveys provide data to estimate transition probabilities between cigarette smoking, e-cigarette use, and dual use of both, facilitating projections of future use and the impact of policies.

**Methods:** We fit a continuous time Markov multi-state model for youth (ages 12-17y) and adults (≥18y) in Waves 1-4.5 of the Population Assessment of Tobacco and Health (PATH) longitudinal survey and estimated smoking and e-cigarette transition frequencies, including initiation, cessation, and relapse. We validated transition frequency results in a microsimulation model by projecting smoking and e-cigarette use prevalence over time.

**Results:** There was more volatility in smoking and e-cigarette use among youth than among adults. For youth never smokers, annual smoking initiation among never/current/former e-cigarette users occurred in 0.4% (95% CI 0.2-0.6%)/8.8% (7.0-10.7%)/3.1% (2.1-4.2%), and current e-cigarette users were more likely to quit e-cigarettes than to initiate smoking (absolute difference in annual probability 46.5%, 38.7-54.2%). For adult current smokers, annual smoking cessation among never/current/former e-cigarette users occurred in 22.6% (20.9-24.3%)/14.5% (11.5-17.4%)/15.1% (12.1-18.2%). For adult current dual users, 14.5% quit smoking and 49.5% quit e-cigarettes annually. For adult former smokers, annual smoking relapse among never/current/former e-cigarette users occurred in 17.7% (15.8-19.6%)/29.3% (23.8-34.7%)/32.8% (27.1-38.6%). Using these transition probabilities in a microsimulation model accurately projected smoking and e-cigarette use prevalence at 12 and 24 months compared to PATH empirical data (root-mean-square error <0.7%).

**Discussion:** PATH Waves 1-4.5 contain sufficient data to generate smoking and e-cigarette use transition frequency estimates for youth and adults in a microsimulation model. E-cigarette use among youth is especially volatile.

**IMPLICATIONS:** This study estimates longitudinal transitions between nine cigarette smoking and e-cigarette use states among both youth and adults. Youth smoking and e-cigarette use was more volatile than that of adults. Smoking initiation, cessation, and relapse differed by e-cigarette use status, but it is not clear that e-cigarette use itself caused these differences. A tobacco and nicotine policy model parametrized with our estimates accurately predicted smoking and e-cigarette use prevalence over two years, enabling future work to project the outcomes and cost-effectiveness of policies targeting smoking and e-cigarette use.

## INTRODUCTION

Electronic cigarettes (e-cigs) are battery-operated nicotine delivery devices that produce an aerosol for inhalation (“vaping”) but do not burn tobacco. There has been a dramatic increase in the use of e-cigs in the last decade in the US, sparking debates about their potential harms and benefits. E-cig use among US high school students rose from 1.5% in 2011 to 27.5% in 2019, raising concern about a youth vaping epidemic and the potential for nicotine addiction or transition to cigarette smoking among those who otherwise might not use tobacco or nicotine products.^1,2^ Conversely, several clinical trials and observational studies have shown that e-cig use can help adults to quit combustible cigarette smoking, and some see e-cig use as a potential harm reduction strategy.^3–6^ Meanwhile, in 2019, in response to the increased youth e-cig prevalence and cases of e-cigarette and vaping related acute lung injury (EVALI), several states and municipalities and the federal government instituted restrictions or bans on e-cig sales.^7–9^ In this context, the net public health impact of e-cig policies, in terms of their impact on youth uptake, smoking cessation or reduction among adults, and direct toxicity, remains unclear. Simulation models provide a useful framework for evaluating downstream outcomes of policies.

To inform simulation models, estimates of transition frequencies between various states of cigarette smoking, e-cig use, and dual use – including initiation, cessation, and relapse – are needed. These estimates, themselves, may also illuminate associations between e-cig use and smoking initiation, cessation, and relapse. There are few published statistical models of tobacco and nicotine product transition frequencies, particularly those that include e-cigs. Published studies model changes in population-level smoking and e-cig use rather than individual-level transitions, combine former smokers and former e-cig users into a single group, do not differentiate current exclusive e-cig users by smoking history, or focus on either adults or on adolescents and young adults but not on both.^10–15^

We sought to estimate individual-level behavioral transitions, among both youth and adults, between each combination of current, former, and never smoking and e-cig use states, to compare frequencies of smoking transitions in the presence and absence of e-cigs, and to determine how accurately these estimates, as inputs to a tobacco and nicotine policy microsimulation model, project smoking and e-cig use and prevalence. If validated, individual-level behavioral transitions can be used in simulation models to project long-term clinical and economic outcomes associated with smoking, e-cig use, and associated regulatory policies.

## METHODS

### Data Source and Statistical Tools

We analyzed transitions in cigarette smoking and e-cig (e-cigarette, vape pen, personal vaporizer and mods, e-cigar, e-pipe, e-hookah or hookah pen) use in the Population Assessment of Tobacco and Health (PATH) Study, a nationally representative longitudinal survey of the US noninstitutionalized population ages 12 years and older.^16–18^ Wave 1 of PATH includes survey responses from 13,500 youth (ages 12-17 years) and 32,000 adults (ages ≥18 years) from September 2013 to December 2014. An additional 7,000 youth ages 9-11 years were identified during the Wave 1 sampling process and were surveyed in subsequent waves after reaching 12 years of age. As of May 2021, survey responses are also available from Wave 2 (October 2014 to October 2015, youth and adults), Wave 3 (October 2015 to October 2016, youth and adults), Wave 4 (December 2016 to January 2018, youth and adults), and Wave 4.5 (December 2017 to December 2018, youth only). All PATH questionnaires evaluate cigarette and e-cig product use, but questions vary between youth and adults and by wave (Figure 1).

**Figure 1:**
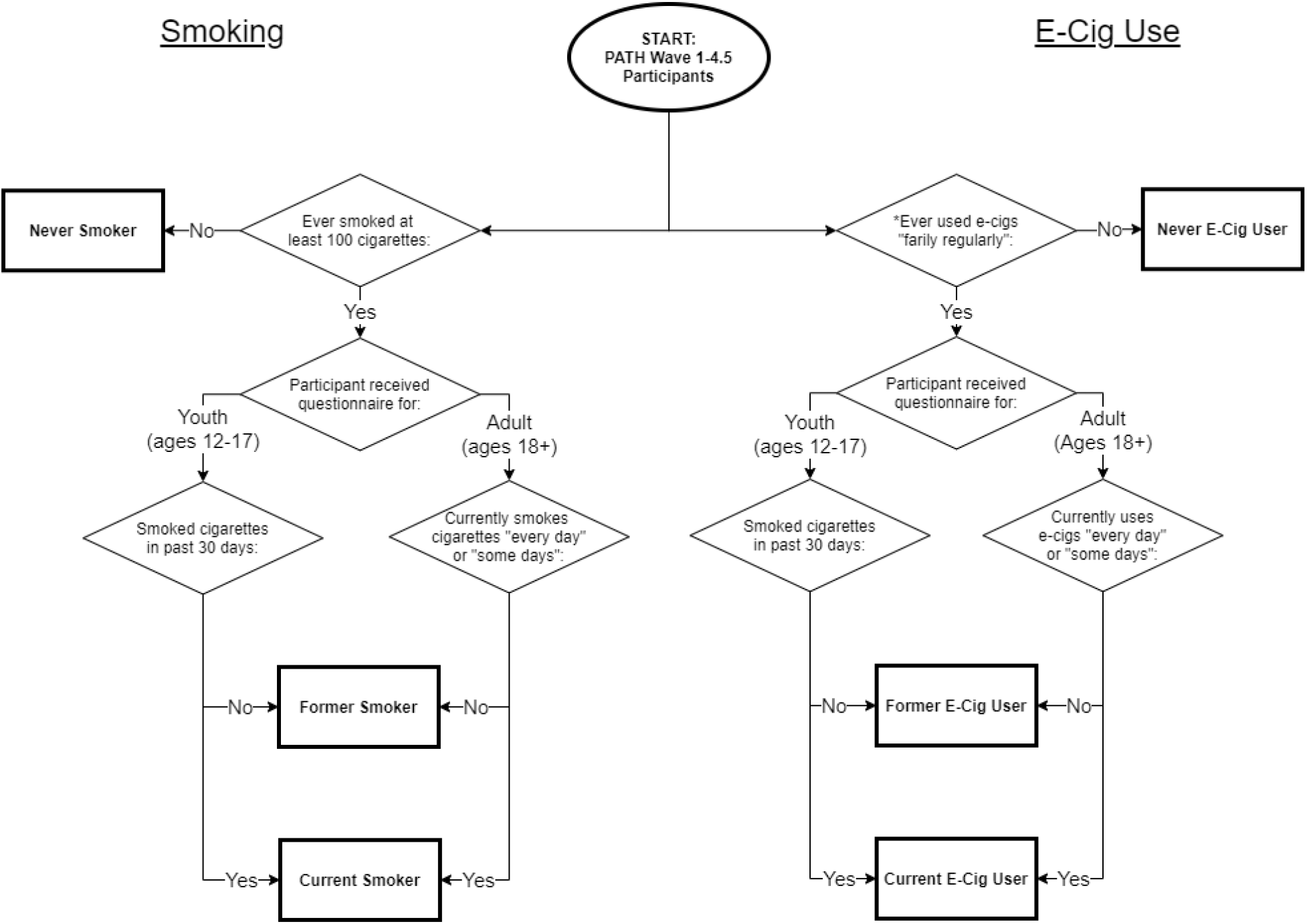
Classification of survey participants as never, former, and current smokers/e-cig users. Population Assessment of Tobacco and Health (PATH) Study Waves 1-4.5 respondents were classified into never, former, and current smoking/e-cig use. A single participant’s classification could differ between the two products: e.g., former smoker/current e-cig user. In youth survey Waves 1-2 and adult survey Wave 1, e-cigs are defined as “electronic-cigarettes.” In all other survey waves, e-cigs are defined as “e-cigarettes, vape pens, personal vaporizers and mods, e-cigars, e-pipes, e-hookah or hookah pens.” *Youth in Wave 1 were asked to attest to ever use instead of ever “fairly regular” use of e-cigs. In our analysis, youth in Wave 1 who attested to never having used e-cigs were classified as never e-cig users. We classified all other youth as “undefined” for Wave 1 e-cig use and excluded them from the analysis in that wave.

The PATH data include weights to adjust for bias introduced by complex survey design and non-response. According to the recommendations of the PATH User Guide, we weighted responses with Wave 4 and 4.5 longitudinal weights.^19^ We conducted analyses in R version 3.6.3 with packages “tidyverse” version 1.3.0 and “msm” version 1.6.8.

### Definitions of Smoking and E-Cig Use Status

We merged data across youth and adult questionnaire responses from Waves 1-4.5 and classified participants as current, former, or never cigarette smokers, and as current, former, or never e-cig users (Figure 1). We considered participants to be established smokers/e-cig users if they had smoked over 100 cigarettes in their lifetime or ever used e-cigs “fairly regularly”, respectively.^20,21^ Among established smokers/e-cig users, we distinguished current from former smoking/e-cig use by past 30-day use for youth and current every-day or some-day use for adults. Participants with no established smoking/e-cig use were classified as never smoker/e-cig users. We excluded survey responses where smoking and e-cig use data were missing, unless participant use status could be determined deductively (i.e. they were classified as a never smoker/never e-cig user in a future wave).

### Continuous Time Markov Multi-State Model

Continuous time Markov multi-state models are suitable for estimating, from longitudinal data, the frequencies at which individuals transition between pre-determined states. These models do not require exact transition times to be observed and allow multiple transitions to occur consecutively between observations.^22^ In this analysis, we defined nine states corresponding to the combinations of: (1) current, former, and never cigarette smoking, and (2) current, former, and never e-cig use. We specified 27 allowable instantaneous state-transitions (Figure 2). We incorporated sex (female or male) and age category at each survey wave (12-17, 18-24, 25-44, ≥45 years) as covariates. We supplemented the msm package with additional code to incorporate survey weights into transition frequency estimates (Supplemental Methods).

**Figure 2.**
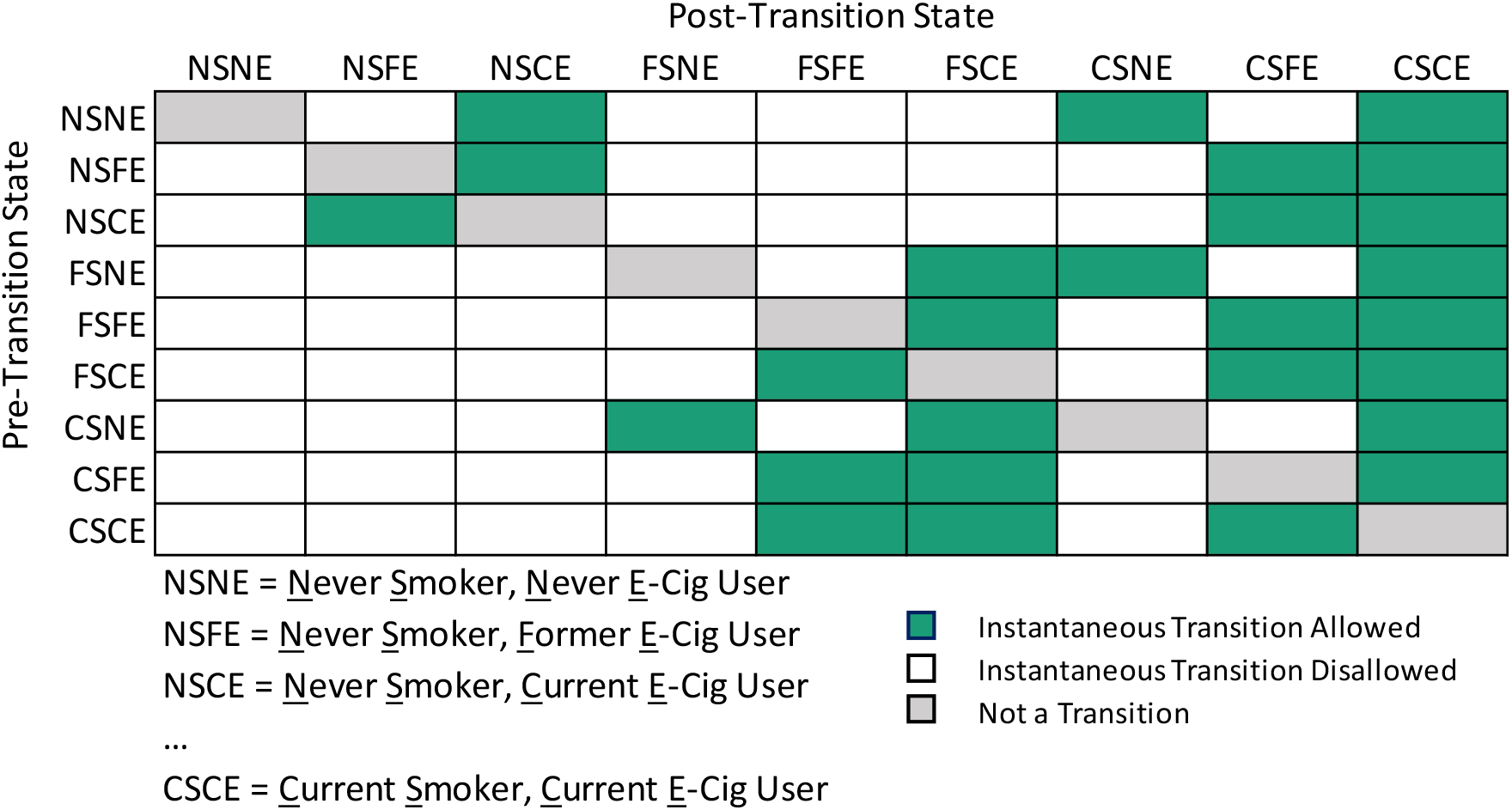
Allowed and disallowed instantaneous transitions in the continuous-time Markov multi-state model of cigarette smoking and e-cig use states. Continuous time Markov multi-state models do not require exact transition times to be observed and allow multiple transitions to occur between observations. They therefore require allowed instantaneous transitions to be specified. Allowed instantaneous transitions from the indicated pre-transition smoking and e-cig use state (row) to the indicated post-transition state (column) are in green. Disallowed instantaneous transitions are in white. Cells in gray are those reflecting staying in the same state.

### Comparisons of Transition Frequencies of High Relevance to Public Health

We leveraged the multi-state model estimates of transition frequencies to examine three associations of particular interest: (1) e-cig use and smoking initiation among youth never smokers, (2) e-cig use and smoking cessation attempts among adult current smokers, and (3) e-cig use and smoking relapse among adult former smokers. We present estimates of the frequencies of transitions between smoking and e-cig use states with three different statistics: transition rates, annual first transition probabilities, and annual cumulative transition probabilities.

The multi-state model directly estimates transition rates, which measure the instantaneous likelihood of each allowable transition. Annual first transition probabilities, derived from transition rates, estimate the proportion of people in a prior state expected to transition to a subsequent state as their first transition of the year. Annual cumulative transition probabilities, also derived from transition rates, estimate the proportion of people in a prior state expected to be in a subsequent state one year later, where some individuals will undergo multiple transitions within the year.^22^ As there is some redundancy in the information conveyed by these statistics, we mainly present annual first transition probabilities and include transition rates and annual cumulative transition probabilities where they may aid in interpretation.

### Internal Validation of Transition Estimates Using A Tobacco and Nicotine Policy Microsimulation Model

We utilized the Simulation of Tobacco and Nicotine Outcomes and Policy (STOP) microsimulation model, now updated to incorporate e-cig use, to assess how accurately a tobacco and nicotine policy model parametrized with our transition frequency estimates could predict prevalence of each of the nine smoking and e-cig use states over time (Supplemental Methods).^23^

We populated STOP’s initial age, sex, and cigarette smoking and e-cig use distributions with the corresponding estimates at PATH Wave 2 (PATH Wave 1 lacked the necessary questions to distinguish between established and experimental e-cig use among youth, Figure 1). We ran the STOP model for 24 months, extracted model-predicted smoking and e-cig use prevalence for the entire population at each month, and compared STOP projected prevalence at 12 months and at 24 months to empirical data from PATH Waves 3 and 4. Participants who were younger than 12 years old at Wave 2, and therefore were not included in the STOP simulated cohort, were excluded from PATH Wave 3 and 4 empirical prevalence estimates. We assessed goodness of model fit via root-mean-square error (RMSE) between projected (STOP model) and empirical (PATH data) prevalence.

### Validation of Time-Variant Relapse

Continuous time Markov multi-state models make the simplifying assumption that transition rates do not depend on the duration spent in the current state (e.g., that a current smoker of one year and a current smoker of 10 years have equal rates of smoking cessation). However, smoking relapse probabilities are known to depend on duration of abstinence.^24,25^ It is plausible that e-cig relapse probabilities follow a similar pattern, given that nicotine is the common addictive drug in both cigarettes and e-cigs. The STOP model allows rates of smoking and e-cig relapse to decay exponentially with respect to the number of months of abstinence (Supplemental Methods). We performed a second, “time-variant relapse” validation in which we utilized this feature over a 24-month time horizon and compared the results to both empirical estimates from PATH and the previously described static relapse validation.

## RESULTS

### Characteristics of Path Data

The weighted PATH survey response rate among adults in Waves 1-4 was 73.5% and among youth in Waves 1-4.5 was 74.6%.^19^ Among those who responded to the survey, the weighted response rate for sex was 99.9%, for age category was >99.9%, and for smoking and e-cig use state was 91.2%. Participants completed a weighted median of 4 survey waves. Supplemental Table S1 includes smoking and e-cig prevalence by age, sex, and wave.

### Multi-State Model: E-Cig Use and Smoking Initiation Among Youth Never Smokers

Smoking initiation was rare among all youth never smokers. The annual first transition probability of smoking initiation among youth never smoker/never e-cig users was 0.4% (95% CI 0.2-0.6%). Never smoker/current e-cig users were more likely to initiate smoking, with a probability of 8.8% (difference of 8.4% vs. never e-cig users, CI 6.5-10.4%). Never smoker/former e-cig users were expected to start smoking with a probability of 3.1%, greater than that of never e-cig users (difference of 2.7%, CI 1.6-3.9%) and less than that of current e-cig users (difference of 5.7%, CI 3.3-8.1%). Notably, annual cumulative transition probabilities show youth never smoker/never e-cig users to be equally likely to transition to current smoking/never e-cig use (0.3%) as they were to transition to current smoking/current or former e-cig use (0.3%; difference of 0.0%, CI 0.0-0.0%; Figure 3).

**Figure 3.**
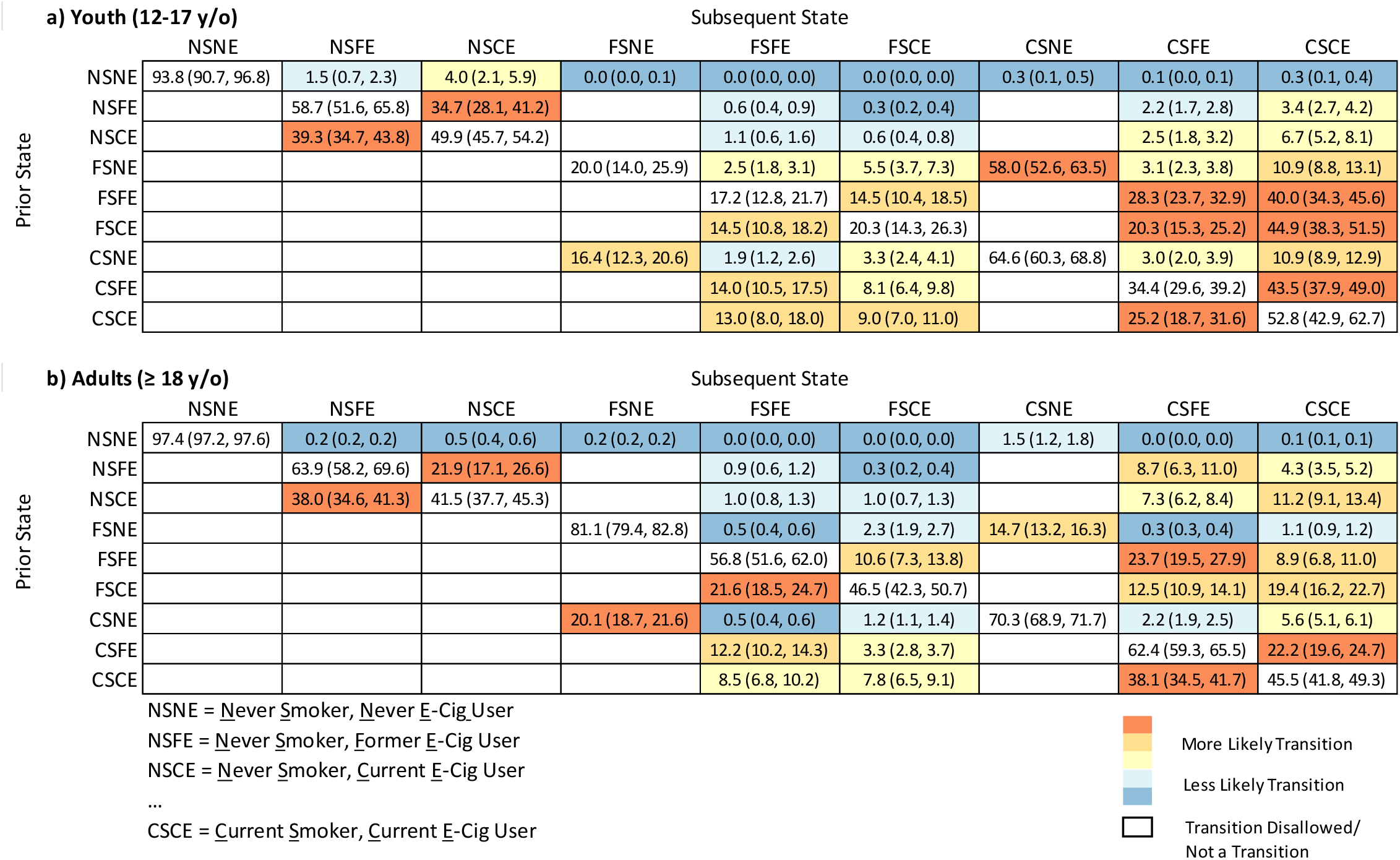
Annual cumulative transition probabilities according to the continuous-time Markov multi-state model for a) youth and b) adults. Data are presented as the probability, among those in the indicated prior state (row), of being in the indicated subsequent state (column) one year later. In parentheses are the 95% confidence intervals. More likely transitions are pictured in orange, while less likely transitions are pictured in blue. Note that participants may experience more than one instantaneous transition in sequence within the year, meaning some transitions that cannot occur instantaneously (e.g., NSNE to FSFE) are allowed as annual cumulative transitions.

Among youth never smokers, e-cig use was substantially more volatile than smoking. Comparing annual first transition probabilities among never smoker/current e-cig users, 55.3% were expected to quit e-cigs and 8.8% were expected to start smoking (difference of 46.5%, CI 38.7-54.2%). Among never smoker/former e-cig users, 53.1% were expected to relapse to exclusive e-cig use and 3.1% were expected to start smoking (difference of 50.0%, CI 39.1-60.9%). Transition rates (Supplemental Table S2) and annual cumulative transition probabilities (Figure 3) show analogous trends.

### Multi-State Model: E-Cig Use and Smoking Cessation Among Adult Current Smokers

The annual first transition probability of smoking cessation among adult current smoker/never e-cig users was 22.6% (CI 20.9-24.3%). Current smoker/current e-cig users were less likely to quit smoking, with a probability of 14.5% (difference of 8.1% vs. never e-cig users, CI 4.7-11.5%). Current smoker/former e-cig users were expected to quit smoking with a probability of 15.1%, less than that of never e-cig users (difference of 7.5%, CI 4.1-10.8%) and similar to that of current e-cig users (difference of 0.6%, CI -4.4 to 5.7%).

Among adult current smokers, e-cig use was more volatile than smoking. Comparing annual first transition probabilities among current smoker/current e-cig users, 49.5% were expected to quit e-cigs and 14.5% were expected to quit smoking (difference of 35.0%, CI 28.2-41.8%). Among current smoker/former e-cig users, 33.6% were expected to restart e-cig use and 15.1% were expected to quit smoking (difference of 18.4%, CI 12.0-24.9%).

### Multi-State Model: E-Cig Use and Smoking Relapse Among Adult Former Smokers

The annual first transition probability of smoking relapse among adult former smoker/never e-cig users was 17.7% (CI 15.8-19.6%). Former smoker/current e-cig users were more likely to relapse to smoking, with a probability of 29.3% (difference of 11.6% vs. never e-cig users, CI 5.8-17.4%). Former smoker/former e-cig users were expected to relapse to smoking with a probability of 32.8%, greater than that of never e-cig users (difference of 15.1%, CI 9.3-21.0%) and similar to that of current e-cig users (difference of 3.6%, CI -5.0 to 12.2%).

Among adult former smokers, e-cig use was less volatile than or equally volatile to smoking. Comparing annual first transition probabilities among former smoker/former e-cig users, 15.0% were expected to restart e-cigs and 32.8% were expected to restart smoking (difference of 17.9%, CI 8.8-27.0%). Among former smoker/current e-cig users, 27.8% were expected to quit e-cigs and 29.3% were expected to restart smoking (difference of 1.5%, CI -7.6 to 10.6%).

### Multi-State Model: Differences in Transitions Between Youth and Adults

Youth smoking and e-cig use was generally more volatile than that of adults. For 17 of the 27 instantaneous transitions between smoking and e-cig use, youth transitioned at a greater rate than adults (multivariable adjusted rate ratio [RR] > 1.2 and p < 0.05). For 4 transitions, youth and adult transition rates were comparable (0.8 < RR < 1.2 and p > 0.05). In 6 transitions, youth rates were lower than those of adults (RR < 0.9 and p < 0.05). This trend persists even when restricting the comparison to youth and young adults (ages 18-24 years, Supplemental Table S2).

Notably, youth former smoker/never e-cig users were substantially more likely to relapse to smoking than were their adult counterparts (annual first transition probabilities 81.1% vs. 17.7%; difference of 63.4%, CI 59.0-67.9%). This was also true of former smoker/former e-cig users, but not of former smoker/current e-cig users (Supplemental Table S2). Youth never smoker/never e-cig users were more likely to initiate e-cig use than their adult counterparts (5.9% vs. 0.8%; difference of 5.0%, CI 2.1-8.0%), but these youth were less likely to start smoking than were adults (0.4% vs. 1.8%; difference of 1.4%, CI 1.3-1.5%). Youth current smoker/current e-cig users were more likely to simultaneously quit smoking and e-cig use than were their adult counterparts (22.9% vs. 4.1%; difference of 18.7%, CI 6.8-30.6%). These differences between youth and adult transition rates translate to analogous differences in annual cumulative transition probabilities (Figure 3).

### Multistate Model: Differences in Transitions Between Men and Women

Smoking and e-cig transitions were similar between men and women. For 7 of the 27 instantaneous transitions between smoking and e-cig use, men transitioned at a greater rate than women (RR > 1.3 and p < 0.05). For 17 transitions, men’s and women’s transition rates were comparable (0.7 < RR < 1.6 and p > 0.05). For 3 transitions, men’s transition rates were lower than women’s (RR < 0.9 and p < 0.05). Male never smoker/never e-cig users were marginally more likely to initiate exclusive smoking than their female counterparts (1.8% vs. 1.3%, difference of 0.5%, CI 0.3-0.8%). These men were also more likely to initiate exclusive e-cig use than were their female counterparts (1.4% vs. 0.8%; difference of 0.6%, CI 0.4-0.8%). Female former smoker/never e-cig users were marginally more likely to relapse to exclusive smoking than were their male counterparts (24.8% vs. 20.3%; difference of 4.4%, CI 0.1-8.7%). Supplemental Table S2 includes all transition rate ratios by sex (female or male).

### Internal Validation of Transition Estimates: Static and Time-Variant Relapse

Root-mean-squared error for projected versus empirical smoking and e-cig use prevalence was less than 0.7% for both the static relapse and time-variant relapse STOP simulations, with no significant difference in goodness of fit between the two (for static relapse: RMSE 0.69%, CI 0.38-0.99%; for time-variant relapse: RMSE 0.65%, CI 0.42-0.87%). The largest contributors of error were, in the static relapse simulation, the proportion of former smoker/never e-cig users at Wave 4 (projected prevalence 20.6% vs. empirical prevalence 22.7%; difference of 2.1%, CI 1.0-3.2%), and, in the time-variant relapse simulation, the proportion of former smoker/never e-cig users at Wave 3 (projected prevalence 20.1% vs. empirical prevalence 21.8%; difference of 1.7%, CI 0.7-2.7%). Empirical estimates of smoking and e-cig use prevalence from PATH were mostly within the margin of error estimated by both static relapse and time-variant relapse simulations (Supplemental Table S3 and Figure S3).

## DISCUSSION

We leveraged a continuous time Markov multi-state model to estimate rates of smoking and e-cig initiation, cessation, and relapse using longitudinal data from PATH Waves 1-4.5, incorporating sex and age as covariates. We validated transition estimates using the STOP microsimulation model, which projected prevalence of e-cig use and cigarette smoking over time that were similar to the empirical prevalence in PATH Wave 3 and Wave 4.

This analysis includes several novel aspects chosen to maximize its utility in parametrizing a tobacco and nicotine policy model. First, our focus on established rather than experimental cigarette smoking and e-cig use is distinct from other work incorporating youth and young adults, and it better aligns with long-term health risks.^11,12,26^ Second, our modeling of all nine combinations of current, former, and never smoking/e-cig use allows future work to ascribe distinct disease incidence and mortality risks (e.g., myocardial infarction, lung cancer) to each use state. Previously published studies of individual-level transitions do not, for example, distinguish never smoker/current or former e-cig users from former smoker/current or former e-cig users, who are likely to experience different morbidity and mortality over time.^10–12,26^ Third, our exploration of relapse rates that decay exponentially with respect to duration of abstinence reflects the reality that most quit attempts result in early relapse and a longer duration of abstinence is associated with a lower probability of relapse.^24,25^ Notably, static relapse and time-variant relapse microsimulations are comparable in their ability to predict PATH Wave 3 and 4 smoking and e-cig use prevalence. Fourth, our simultaneous analysis of both youth and adults is also novel. We found that smoking and e-cig use was generally more volatile among youth than adults, even when restricting the comparison to youth versus young adults ages 18-24 years. Quantifying such differences facilitates future analysis to capture any differential impact among youth and adults of policy interventions around cigarette smoking and e-cig use.^10–12,15^

This study cannot determine causal relationships. However, it adds nuance to the evidence of the associations between e-cig use and smoking transitions. Among both youth and adult never smokers, there was a significant positive association between current or former e-cig use and smoking initiation, consistent with the findings of previous multi-state models of smoking and e-cig use.^10–12^ In contrast to these prior studies, we separated never smoker/current and former e-cig users from former smoker/current and former e-cig users, and could therefore limit the confounding associated with smoking history. We found that youth never smoker/never e-cig users were as likely annually to initiate smoking directly as they were to initiate e-cig use followed by smoking, and that never smoker/current and former e-cig users were substantially more likely to change their e-cig use status than they were to initiate smoking. It remains unclear if or to what degree e-cig use may cause subsequent smoking among cigarette-naïve youth, but we found that e-cig use was not the most influential factor driving youth to initiate smoking.

Youth e-cig uptake itself, however, is noteworthy. We found that youth never smoker/never e-cig users were less likely than their adult counterparts to start smoking but more likely to start using e-cigs. Youth current e-cig users were very likely to quit, but equally likely to subsequently restart e-cig use even if they did quit, a dynamic that can only be captured by longitudinal data such as those in the PATH Study. Further study is warranted to investigate if this instability is indicative of casual use that is dependent upon social context (e.g., students who start and stop e-cig use based on the behavior of their peers) or is indicative of nicotine addiction and difficulty maintaining abstinence after cessation attempts.

We found significant negative associations between adult e-cig use and the likelihood of smoking cessation and positive associations between adult e-cig use and the likelihood of smoking relapse. A possible interpretation is that e-cig use may interfere with abstinence from smoking. However, this is not consistent with our finding that the quit and relapse rates of current or former smoker/current e-cig users were comparable to those of current or former smoker/former e-cig users; among the latter group, there is no apparent ongoing e-cig use to explain interference with smoking quit attempts. More likely, current and former smokers with a history of failed quit attempts or barriers to quitting turn to e-cigs in an effort to reduce or quit smoking. As the PATH Study collects additional waves of data, future models may incorporate the number of previous quit attempts and other sociodemographic variables as covariates to adjust for this confounding.

The magnitude and frequency of significant differences between transition rates was much greater when comparing youth and adults than when comparing men and women, and most of the significant differences between men and women occurred among infrequent transitions. Future modeling work seeking to limit the number of covariates may benefit from prioritizing the incorporation of age over sex.

This work has several limitations. The majority of observations in PATH Waves 1-4.5 predate the rapid rise of pod-based e-cigs such as JUUL in late 2017 and 2018, the EVALI reports in 2019-2020, and the COVID-19 pandemic that emerged in the US in 2020. Considering the higher nicotine content and more rapid nicotine absorption of JUUL products relative to other e-cigs, and the collateral consequences of the EVALI outbreak and the COVID-19 pandemic, it is plausible that the dynamics of smoking and e-cig use may have changed since 2018. When future waves of PATH are released, transition parameter estimates may be updated accordingly. In internal validation, both STOP simulations were less successful at predicting the prevalence of former smoker/never e-cig users than of other use states, suggesting that our estimates of smoking cessation and relapse would benefit from further refining. When future waves of PATH are released, there may be sufficient data to model time-variant smoking relapse using a semi-Markov multi-state model (which includes unobserved sub-states of former smokers representing high versus low relapse likelihood), rather than relying on external smoking relapse data.^22,23^ Our estimates account for sex and age but not for race, socio-economic status, level of nicotine dependence, and exposure to product advertising, which may be associated with differences in cigarette smoking and e-cig use behaviors. Our inclusion of more states and transitions than previous models made the computational intensity of including such variables infeasible. Nonetheless, our results can be integrated with race/ethnicity, education, and income hazard ratios estimated by others to project smoking and e-cig use stratified by these characteristics.^10^

Our results indicate that PATH Waves 1-4.5 contain sufficient observations to parametrize a continuous-time Markov multi-state model with nine cigarette smoking and e-cig use states, 27 allowable instantaneous transitions, two sex categories, and four age categories. Model-estimated transition rates differ substantially by age and transition type, and a tobacco and nicotine policy model parametrized with these estimates predicted smoking and e-cig use prevalence over a 2-year time horizon with RMSE <0.7%. Tobacco and nicotine product use, especially e-cig use, was more volatile among youth than among adults. In future work, simulation models can be populated with these estimates along with additional data on smoking and e-cig use-stratified mortality, noncommunicable disease incidence, quality of life, and health care costs to predict the outcomes and cost-effectiveness of proposed policy interventions targeting smoking and e-cig use.^23^

## Data Availability

The Population Assessment of Tobacco and Health (PATH) data underlying this article are available from the Interuniversity Consortium for Political and Social Research at https://dx.doi.org/10.3886/SERIES606. Our code is available upon request to the corresponding author.

## FUNDING

This work was supported by awards from the National Institute on Drug Abuse of the National Institutes of Health (NIH) (K01 DA042687) and the National Cancer Institute of the NIH and Food and Drug Administration Center for Tobacco Products (U54 CA229974). The funding sources had no role in the study design, data collection, data analysis, data interpretation, writing of the manuscript, or in the decision to submit the manuscript for publication. The content is solely the responsibility of the authors and does not necessarily represent the official views of the National Institutes of Health or the Food and Drug Administration.

## DECLARATION OF INTERESTS

Dr. Rigotti and Dr. Reddy receive royalties from UpToDate, Inc. Dr. Rigotti has received a research grant from and consulted with Achieve Life Sciences regarding an investigational smoking cessation drug. The other authors have no conflicts of interest to declare.

## Notes

### Author Declarations

Approval was obtained from the National Addiction & HIV Data Archive Program (NAHDAP) to access the Population Assessment of Tobacco and Health (PATH) Study data. The Mass General Brigham Institutional Review Board approved the project (Protocol # 2019P001772).

## REFERENCES

1. Gentzke AS. Vital Signs: Tobacco Product Use Among Middle and High School Students — United States, 2011–2018. MMWR Morb Mortal Wkly Rep. 2019;68. doi:10.15585/mmwr.mm6806e1

2. Gentzke AS, Wang TW, Jamal A, et al. Tobacco Product Use Among Middle and High School Students — United States, 2020. MMWR Morb Mortal Wkly Rep. 2020;69(50):8.

3. Hartmann-Boyce J, McRobbie H, Lindson N, et al. Electronic cigarettes for smoking cessation. Cochrane Database Syst Rev. 2020;10:CD010216. doi:10.1002/14651858.CD010216.pub4

4. Hajek P, Phillips-Waller A, Przulj D, et al. A Randomized Trial of E-Cigarettes versus Nicotine-Replacement Therapy. N Engl J Med. 2019;380(7):629–637. doi:10.1056/NEJMoa1808779

5. Eisenberg MJ, Hébert-Losier A, Windle SB, et al. Effect of e-Cigarettes Plus Counseling vs Counseling Alone on Smoking Cessation: A Randomized Clinical Trial. JAMA. 2020;324(18):1844–1854. doi:10.1001/jama.2020.18889

6. Kalkhoran S, Chang Y, Rigotti NA. Electronic Cigarette Use and Cigarette Abstinence Over Two Years among U.S. Smokers in the Population Assessment of Tobacco and Health Study. Nicotine Tob Res. Published online July 11, 2019. doi:10.1093/ntr/ntz114

7. 2019 Tobacco Control Law | Mass.gov. Accessed June 3, 2021. https://www.mass.gov/guides/2019-tobacco-control-law

8. Office of the Commissioner. FDA finalizes enforcement policy on unauthorized flavored cartridge-based e-cigarettes that appeal to children, including fruit and mint. FDA. Published March 24, 2020. Accessed June 3, 2021. https://www.fda.gov/news-events/press-announcements/fda-finalizes-enforcement-policy-unauthorized-flavored-cartridge-based-e-cigarettes-appeal-children

9. Sindelar JL. Regulating Vaping — Policies, Possibilities, and Perils. N Engl J Med. 2020;382(20):e54. doi:10.1056/NEJMp1917065

10. Brouwer AF, Jeon J, Hirschtick JL, et al. Transitions between cigarette, ENDS and dual use in adults in the PATH study (waves 1–4): multistate transition modelling accounting for complex survey design. Tob Control. Published online November 15, 2020. doi:10.1136/tobaccocontrol-2020-055967

11. Hair EC, Romberg AR, Niaura R, et al. Longitudinal Tobacco Use Transitions Among Adolescents and Young Adults: Nicotine Tob Res. 2019;21(4):458–468. doi:10.1093/ntr/ntx285

12. Niaura R, Rich I, Johnson AL, et al. Young Adult Tobacco and E-cigarette Use Transitions: Examining Stability Using Multistate Modeling. Nicotine Tob Res. 2020;22(5):647–654. doi:10.1093/ntr/ntz030

13. Apelberg BJ, Feirman SP, Salazar E, et al. Potential Public Health Effects of Reducing Nicotine Levels in Cigarettes in the United States. N Engl J Med. 2018;378(18):1725–1733. doi:10.1056/NEJMsr1714617

14. Vugrin ED, Rostron BL, Verzi SJ, et al. Modeling the Potential Effects of New Tobacco Products and Policies: A Dynamic Population Model for Multiple Product Use and Harm. PLoS ONE. 2015;10(3):e0121008. doi:10.1371/journal.pone.0121008

15. Stanton CA, Bansal-Travers M, Johnson AL, et al. Longitudinal e-cigarette and cigarette use among US youth in the PATH Study (2013-2015). J Natl Cancer Inst. Published online January 25, 2019. doi:10.1093/jnci/djz006

16. United States Department Of Health And Human Services. National Institutes of Health. National Institute on Drug Abuse. Population Assessment of Tobacco and Health (PATH) Series. Published online 2016. doi:10.3886/SERIES606

17. Kasza KA, Ambrose BK, Conway KP, et al. Tobacco-Product Use by Adults and Youths in the United States in 2013 and 2014. N Engl J Med. 2017;376(4):342–353. doi:10.1056/NEJMsa1607538

18. Hyland A, Ambrose BK, Conway KP, et al. Design and methods of the Population Assessment of Tobacco and Health (PATH) Study. Tob Control. 2017;26(4):371–378. doi:10.1136/tobaccocontrol-2016-052934

19. Westat Corporation. Population Assessment of Tobacco and Health (PATH) Study [United ICPSR 37786 States] Special Collection Public-Use Files. Accessed May 12, 2021. https://www.icpsr.umich.edu/web/NAHDAP/studies/37786/datadocumentation

20. NHIS - Adult Tobacco Use - Glossary. Published May 10, 2019. Accessed May 12, 2021. https://www.cdc.gov/nchs/nhis/tobacco/tobacco_glossary.htm

21. Levy DT, Yuan Z, Li Y, Mays D, Sanchez-Romero LM. An Examination of the Variation in Estimates of E-Cigarette Prevalence among U.S. Adults. Int J Environ Res Public Health. 2019;16(17). doi:10.3390/ijerph16173164

22. Jackson C. Multi-state modelling with R: the msm package. https://cran.r-project.org/web/packages/msm/vignettes/msm-manual.pdf

23. Reddy KP, Bulteel AJB, Levy DE, et al. Novel microsimulation model of tobacco use behaviours and outcomes: calibration and validation in a US population. BMJ Open. 2020;10(5). doi:10.1136/bmjopen-2019-032579

24. Stapleton JA, West R. A direct method and ICER tables for the estimation of the cost-effectiveness of smoking cessation interventions in general populations: application to a new cytisine trial and other examples. Nicotine Tob Res. 2012;14(4):463–471. doi:10.1093/ntr/ntr236

25. Yong H-H, Borland R, Cummings KM, Partos T. Do predictors of smoking relapse change as a function of duration of abstinence? Findings from the United States, Canada, United Kingdom and Australia. Addiction. 2018;113(7):1295–1304. doi:10.1111/add.14182

26. Thun MJ, Carter BD, Feskanich D, et al. 50-Year Trends in Smoking-Related Mortality in the United States. N Engl J Med. 2013;368(4):351–364. doi:10.1056/NEJMsa1211127

