## Supplement for "Longitudinal Transitions in Initiation, Cessation, and Relapse of Smoking and E-Cigarette Use Among US Youth and Adults"

### SUPPLEMENTAL METHODS

#### Adding Survey Weights to The MSM Package

By default, the R "msm" package estimates transition rates via maximum likelihood estimation.[1] An optimization algorithm accepts a starting set of transition rate estimates, calculates the model's log-likelihood for those estimates, and iteratively adjusts the estimates and recalculates the log-likelihood until arriving at a solution: a set of transition rate estimates that produce the maximum log-likelihood value.[2] Annual transition probabilities are subsequently calculated from final transition rates.[1] Our R code for *weighted* maximum likelihood estimation pauses the optimization algorithm during every iteration, calculates each survey participant's individual contribution to the model log-likelihood, multiplies by the participants' longitudinal survey weights, and sums the result to estimate the overall model log-likelihood for the transition rate estimates. We estimate weighted transition probabilities using the default msm package function, substituting weighted transition rates for the default unweighted rates.

Conceptually, this is identical to a participant's responses being duplicated in the survey $w_i$ times, where $w_i$ is the value of that participant's longitudinal survey weight. This approach is appropriate for calculating point-estimates of transition rates, but not for calculating their variances. We calculated weighted variances by fitting 100 separate models using each of the 100 PATH-provided replicate-weights and following *Equation 1)*, as recommended by the PATH User Guide.[3]

$$v(\hat{\theta}) = 0.020408 * \sum_{g=1}^{100}\left(\hat{\theta}_{(g)} - \hat{\theta}\right)^2 \qquad \text{Equation 1)}$$

where $\theta$ is the statistic of interest: in this case a transition rate, an annual first or cumulative transition probability, a rate ratio, or a difference in annual first or cumulative transition probabilities; $\hat{\theta}$ is the estimate of $\theta$ calculated using the full-sample longitudinal survey weight; $v(\hat{\theta})$ is the variance of $\hat{\theta}$; and $\hat{\theta}_{(g)}$ is the estimate of $\theta$ using the $g$-th replicate weight.

To reduce computational requirements, we used unweighted transition rates as estimate starting values and limited the optimization algorithm to 25,000 iterations. Supplemental Figure S1 shows the convergence of transition rate estimates with respect to iteration of the weighted maximum likelihood algorithm.

**Details of the STOP Model**

We expanded the Simulation of Tobacco and Nicotine Outcomes and Policy (STOP) microsimulation model to include the same nine smoking and e-cig use states and 27 state-transitions that were specified in the continuous time Markov multi-state model.[4] STOP transitions, however, are divided into three types: start, quit, and relapse (Supplemental Figure S2). Monthly first transition probabilities are specified for each of the 27 state-transitions, as the STOP model only allows one transition per month.[4] Start and quit probabilities are further stratified by age and sex, and relapse probabilities are further stratified by age only.

STOP is stochastic—it uses computer random number generation to simulate a cohort of patients passing through simulated health states with varying probabilities—and uses a monthly time step. It contains additional modules not utilized in this validation that incorporate chronic disease incidence and complications, mortality, and smoking and e-cig policy interventions. STOP does not model birth or other migration into the cohort, so comparison targets needed to come from a longitudinal study. Due to limited availability of external longitudinal data on smoking and e-cig use, we validated STOP model output with the PATH data.

When the time-variant relapse module of STOP is activated, relapse rates decay exponentially over time according to the equation $Ce^{-0.33t}$, where $t$ is the number of months since the participant's last quit attempt and $C$ is the initial relapse rate.[4] The value of $C$ is set such that the overall probability of relapse in the first year after a cessation attempt is equal between static relapse and time-variant relapse simulations.

### SUPPLEMENTAL REFERENCES

1. Jackson C. Multi-state modelling with R: the msm package. https://cran.r-project.org/web/packages/msm/vignettes/msm-manual.pdf

2. Myung IJ. Tutorial on maximum likelihood estimation. *Journal of Mathematical Psychology*. 2003;47(1):90-100. doi:10.1016/S0022-2496(02)00028-7

3. Westat Corporation. Population Assessment of Tobacco and Health (PATH) Study [United ICPSR 37786 States] Special Collection Public-Use Files. Accessed May 12, 2021. https://www.icpsr.umich.edu/web/NAHDAP/studies/37786/datadocumentation

4. Reddy KP, Bulteel AJB, Levy DE, et al. Novel microsimulation model of tobacco use behaviours and outcomes: calibration and validation in a US population. *BMJ Open*. 2020;10(5). doi:10.1136/bmjopen-2019-032579

### SUPPLEMENTAL TABLES AND FIGURES

**Supplemental Table S1.** Included PATH participants by survey wave, age, sex, and smoking and e-cigarette use state, n (weighted %).

| | | Smoking and E-Cig Use State | | | | | | | | |
|---|---|---|---|---|---|---|---|---|---|---|
| Wave | Covariate | CSCE | CSFE | CSNE | FSCE | FSFE | FSNE | NSCE | NSFE | NSNE |
| 1 | *Age* | | | | | | | | | |
| | 12 to 17 years old | 0* | 0* | 131 (1.8%) | 0* | 0* | 17 (0.2%) | 0* | 0* | 7,679 (97.9%) |
| | 18 to 24 years old | 168 (2.3%) | 81 (1.1%) | 1,211 (16.3%) | 37 (0.5%) | 29 (0.4%) | 247 (3.9%) | 78 (1.1%) | 31 (0.4%) | 3,755 (74.0%) |
| | 25 to 44 years old | 295 (2.3%) | 115 (0.9%) | 2,589 (20.1%) | 104 (0.8%) | 56 (0.4%) | 991 (13.5%) | 33 (0.3%) | 11 (0.1%) | 3,131 (61.6%) |
| | 45 or more years old | 174 (0.9%) | 67 (0.3%) | 2,567 (13.9%) | 94 (0.5%) | 36 (0.2%) | 1,712 (28.6%) | 33 (0.1%) | 12 (0.1%) | 3,026 (55.4%) |
| | *Sex* | | | | | | | | | |
| | Female | 319 (1.3%) | 132 (0.5%) | 3,326 (13.4%) | 134 (0.6%) | 64 (0.3%) | 1,375 (16.9%) | 71 (0.2%) | 27 (0.1%) | 9,174 (66.7%) |
| | Male | 318 (1.6%) | 131 (0.7%) | 3,172 (17.1%) | 101 (0.6%) | 57 (0.3%) | 1,592 (19.9%) | 73 (0.3%) | 27 (0.1%) | 8,417 (59.4%) |
| 2 | *Age* | | | | | | | | | |
| | 12 to 17 years old | 34 (0.4%) | 9 (0.1%) | 84 (1.1%) | 0 | 7 (0.1%) | 27 (0.4%) | 79 (1.1%) | 58 (0.7%) | 7,894 (96.1%) |
| | 18 to 24 years old | 259 (3.4%) | 182 (2.5%) | 988 (13.0%) | 56 (0.8%) | 63 (0.9%) | 302 (4.3%) | 154 (2.1%) | 113 (1.6%) | 4,193 (71.4%) |
| | 25 to 44 years old | 415 (3.2%) | 336 (2.5%) | 2,370 (18.3%) | 189 (1.5%) | 108 (0.8%) | 1,168 (14.4%) | 37 (0.3%) | 35 (0.3%) | 3,156 (58.9%) |
| | 45 or more years old | 240 (1.2%) | 210 (1.1%) | 2,591 (13.9%) | 121 (0.7%) | 76 (0.4%) | 2,001 (30.8%) | 18 (0.1%) | 8 (0.0%) | 2,770 (51.9%) |
| | *Sex* | | | | | | | | | |
| | Female | 467 (1.8%) | 385 (1.4%) | 3,087 (12.5%) | 175 (0.7%) | 124 (0.5%) | 1,634 (18.3%) | 120 (0.4%) | 82 (0.2%) | 9,450 (64.1%) |
| | Male | 481 (2.3%) | 352 (1.8%) | 2,946 (15.8%) | 191 (1.1%) | 130 (0.6%) | 1,864 (21.4%) | 168 (0.6%) | 132 (0.5%) | 8,563 (55.9%) |
| 3 | *Age* | | | | | | | | | |
| | 12 to 17 years old | 30 (0.4%) | 15 (0.2%) | 46 (0.6%) | 4 (0.1%) | 12 (0.1%) | 17 (0.2%) | 97 (1.2%) | 74 (0.9%) | 7,939 (96.2%) |
| | 18 to 24 years old | 239 (3.6%) | 262 (4.1%) | 634 (9.4%) | 73 (1.2%) | 97 (1.5%) | 215 (3.7%) | 185 (2.6%) | 199 (2.9%) | 4,125 (71.1%) |
| | 25 to 44 years old | 363 (3.2%) | 559 (4.7%) | 1,784 (15.7%) | 189 (1.7%) | 227 (1.9%) | 1,043 (14.7%) | 44 (0.3%) | 67 (0.5%) | 2,745 (57.3%) |
| | 45 or more years old | 212 (1.2%) | 340 (1.8%) | 2,117 (12.9%) | 152 (1.0%) | 111 (0.6%) | 1,961 (34.3%) | 11 (0.1%) | 12 (0.1%) | 2,216 (48.1%) |
| | *Sex* | | | | | | | | | |
| | Female | 416 (1.8%) | 602 (2.4%) | 2,370 (11.2%) | 198 (1.0%) | 202 (0.9%) | 1,472 (19.5%) | 130 (0.4%) | 152 (0.4%) | 8,859 (62.4%) |
| | Male | 428 (2.3%) | 574 (3.2%) | 2,211 (13.3%) | 220 (1.3%) | 245 (1.3%) | 1,764 (23.7%) | 207 (0.7%) | 200 (0.8%) | 8,166 (53.4%) |

**Supplemental Table S1, Continued.**

| Wave | Covariate | Smoking and E-Cig Use State | | | | | | | | |
|---|---|---|---|---|---|---|---|---|---|---|
| | | CSCE | CSFE | CSNE | FSCE | FSFE | FSNE | NSCE | NSFE | NSNE |
| 4 | *Age* | | | | | | | | | |
| | 12 to 17 years old | 14 (0.2%) | 12 (0.2%) | 23 (0.4%) | 4 (0.0%) | 4 (0.1%) | 12 (0.2%) | 84 (1.1%) | 78 (1.0%) | 7,711 (97.0%) |
| | 18 to 24 years old | 242 (3.3%) | 294 (4.3%) | 554 (7.5%) | 88 (1.4%) | 121 (1.8%) | 218 (3.3%) | 203 (2.7%) | 286 (3.8%) | 4,986 (71.9%) |
| | 25 to 44 years old | 381 (3.1%) | 747 (6.1%) | 1,674 (14.3%) | 184 (1.7%) | 282 (2.4%) | 1,127 (14.8%) | 47 (0.4%) | 96 (0.7%) | 2,913 (56.5%) |
| | 45 or more years old | 200 (1.1%) | 443 (2.3%) | 2,079 (12.3%) | 153 (0.9%) | 161 (1.0%) | 2,095 (35.0%) | 10 (0.1%) | 16 (0.1%) | 2,244 (47.1%) |
| | *Sex* | | | | | | | | | |
| | Female | 408 (1.7%) | 761 (3.1%) | 2,230 (10.3%) | 197 (0.9%) | 263 (1.2%) | 1,586 (20.2%) | 127 (0.4%) | 210 (0.6%) | 9,265 (61.7%) |
| | Male | 429 (2.1%) | 735 (4.0%) | 2,100 (12.4%) | 232 (1.3%) | 305 (1.7%) | 1,866 (24.2%) | 217 (0.8%) | 266 (0.9%) | 8,589 (52.6%) |
| 4.5 | *Sex* | | | | | | | | | |
| | Female | 14 (0.4%) | 4 (0.1%) | 19 (0.5%) | 4 (0.1%) | 0 | 5 (0.2%) | 95 (2.6%) | 21 (0.6%) | 3,605 (95.4%) |
| | Male | 15 (0.4%) | 13 (0.3%) | 18 (0.5%) | 3 (0.1%) | 4 (0.2%) | 7 (0.2%) | 109 (2.8%) | 52 (1.3%) | 3,820 (94.2%) |

NSNE = Never Smoker, Never E-Cig User
NSFE = Never Smoker, Former E-Cig User
NSCE = Never Smoker, Current E-Cig User
…
CSCE = Current Smoker, Current E-Cig User

PATH = Population Assessment of Tobacco and Health Study.
*Current and former e-cig use could not be ascertained among Wave 1 youth due to limitations in the PATH questionnaire

**Supplemental Table S2.** Continuous time Markov model-estimated baseline transition rates and adjusted rate ratios.

| Transition | Transition Rate: Baseline (12 to 17 years old, Female), Rate per 100 Person-Years (95% CI) | Transition Rate Ratio: 18 to 24 years old Ratio (95% CI) | Transition Rate Ratio: 25 to 44 years old Ratio (95% CI) | Transition Rate Ratio: ≥ 45 years old Ratio (95% CI) | Transition Rate Ratio: Male Ratio (95% CI) |
|---|---|---|---|---|---|
| NSNE to NSCE | 4.57 (0.03, 0.08) | 1.19 (0.86, 1.64) | **0.17 (0.09, 0.31)** | **0.07 (0.03, 0.16)** | **1.79 (1.54, 2.08)** |
| NSNE to CSNE | 0.33 (0.00, 0.01) | **4.14 (2.56, 6.67)** | **3.67 (2.27, 5.93)** | **5.46 (3.46, 8.60)** | **1.44 (1.24, 1.67)** |
| NSNE to CSCE | 0.00 (0.00, 0.00) | 1.01 (0.85, 1.21) | 0.82 (0.64, 1.05) | **2.42 (1.65, 3.55)** | **1.53 (1.23, 1.90)** |
| NSFE to NSCE | 79.47 (0.55, 1.15) | **0.48 (0.36, 0.64)** | **0.38 (0.24, 0.58)** | 0.89 (0.65, 1.21) | 0.96 (0.71, 1.31) |
| NSFE to CSFE | 4.35 (0.03, 0.06) | 1.09 (0.71, 1.67) | **1.40 (1.09, 1.80)** | **5.95 (4.24, 8.35)** | 1.10 (0.86, 1.41) |
| NSFE to CSCE | 0.03 (0.00, 0.81) | **0.81 (0.66, 0.99)** | **0.54 (0.44, 0.67)** | **1.90 (1.56, 2.31)** | 1.11 (0.88, 1.41) |
| NSCE to NSFE | 93.20 (0.76, 1.15) | 1.12 (0.92, 1.38) | 1.07 (0.82, 1.39) | 0.82 (0.61, 1.09) | 0.90 (0.75, 1.07) |
| NSCE to CSFE | 0.01 (0.00, 0.18) | **0.65 (0.50, 0.85)** | **1.89 (1.46, 2.43)** | **3.49 (2.68, 4.56)** | 0.99 (0.65, 1.49) |
| NSCE to CSCE | 12.00 (0.09, 0.15) | 1.23 (0.90, 1.69) | 1.10 (0.83, 1.45) | **3.07 (2.27, 4.16)** | **1.40 (1.13, 1.72)** |
| FSNE to FSCE | 25.26 (0.19, 0.34) | 1.00 (0.74, 1.36) | **0.21 (0.16, 0.27)** | **0.05 (0.03, 0.07)** | 1.52 (0.96, 2.40) |
| FSNE to CSNE | 249.81 (1.92, 3.24) | **0.57 (0.47, 0.69)** | **0.15 (0.13, 0.19)** | **0.04 (0.03, 0.05)** | **0.81 (0.65, 1.00)** |
| FSNE to CSCE | 0.13 (0.00, 0.90) | **0.48 (0.39, 0.59)** | **0.51 (0.42, 0.62)** | **0.47 (0.37, 0.60)** | **0.21 (0.15, 0.30)** |
| FSFE to FSCE | 104.65 (0.70, 1.56) | **0.34 (0.26, 0.44)** | **0.30 (0.23, 0.38)** | **0.12 (0.09, 0.16)** | 1.04 (0.76, 1.42) |
| FSFE to CSFE | 155.29 (1.14, 2.12) | **0.44 (0.36, 0.54)** | **0.27 (0.21, 0.36)** | **0.24 (0.17, 0.33)** | 0.89 (0.66, 1.21) |
| FSFE to CSCE | 25.25 (0.11, 0.58) | 1.25 (0.88, 1.78) | **0.15 (0.12, 0.19)** | **0.05 (0.04, 0.06)** | **3.91 (3.03, 5.05)** |
| FSCE to FSFE | 71.43 (0.54, 0.95) | 1.21 (0.92, 1.58) | **0.77 (0.61, 0.96)** | **0.42 (0.34, 0.53)** | 0.93 (0.72, 1.21) |
| FSCE to CSFE | 10.12 (0.06, 0.18) | **13.88 (11.39, 16.92)** | **0.18 (0.14, 0.23)** | **0.16 (0.09, 0.29)** | **0.14 (0.10, 0.20)** |
| FSCE to CSCE | 125.53 (0.90, 1.75) | **0.49 (0.38, 0.62)** | **0.38 (0.28, 0.52)** | **0.23 (0.17, 0.30)** | 1.26 (0.92, 1.74) |
| CSNE to FSNE | 65.70 (0.53, 0.81) | **0.56 (0.48, 0.66)** | **0.38 (0.32, 0.44)** | **0.43 (0.36, 0.51)** | 0.94 (0.82, 1.08) |
| CSNE to FSCE | 0.86 (0.00, 0.01) | 1.14 (0.91, 1.43) | 1.10 (0.81, 1.49) | **0.58 (0.46, 0.75)** | **1.69 (1.29, 2.21)** |
| CSNE to CSCE | 18.13 (0.15, 0.22) | **1.23 (1.07, 1.42)** | **0.75 (0.66, 0.86)** | **0.36 (0.31, 0.42)** | 1.01 (0.89, 1.15) |
| CSFE to FSFE | 55.55 (0.42, 0.73) | **0.55 (0.43, 0.70)** | **0.35 (0.27, 0.46)** | **0.29 (0.23, 0.36)** | 1.14 (0.87, 1.49) |
| CSFE to FSCE | 0.61 (0.00, 0.02) | **0.37 (0.28, 0.47)** | 0.82 (0.61, 1.11) | **2.70 (2.02, 3.61)** | 1.09 (0.87, 1.37) |
| CSFE to CSCE | 139.16 (1.11, 1.74) | **0.53 (0.43, 0.65)** | **0.44 (0.35, 0.55)** | **0.30 (0.24, 0.37)** | 0.80 (0.61, 1.03) |
| CSCE to FSFE | 22.53 (0.10, 0.51) | **0.27 (0.18, 0.39)** | **0.07 (0.06, 0.10)** | **0.26 (0.15, 0.46)** | **3.11 (2.34, 4.14)** |
| CSCE to FSCE | 15.84 (0.12, 0.21) | **2.05 (1.64, 2.56)** | 1.16 (0.90, 1.50) | **0.75 (0.59, 0.95)** | 1.09 (0.84, 1.41) |
| CSCE to CSFE | 64.03 (0.42, 0.98) | 1.26 (0.96, 1.65) | 1.29 (0.93, 1.80) | 1.23 (0.88, 1.71) | 0.97 (0.82, 1.13) |

Transition rate ratios, adjusted by sex (female or male) and age category (12-17, 18-24, 25-44, ≥45 years), are juxtaposed against baseline transition rates for female youth (ages 12-17 years). To estimate a transition rate for any subgroup, the baseline rate is multiplied by the corresponding subgroup rate ratio(s). E.g., men ages 25-44 transition from NSNE to NSCE at a rate of 4.57*0.17*1.79 = 1.39 per 100 person-years.

NSNE = Never Smoker, Never E-Cig User
NSFE = Never Smoker, Former E-Cig User
NSCE = Never Smoker, Current E-Cig User
…
CSCE = Current Smoker, Current E-Cig User

**Bold** indicates $p < 0.05$

**Supplemental Table S3**. Validation of transition frequency estimates in the Simulation of Tobacco and Nicotine Outcomes and Policy (STOP) microsimulation model.

| Wave | Smoking and E-Cig Use State | Static Relapse Projection | Time-Variant Relapse Projection | Empirical Estimate | Static Relapse Error: Estimate (95% CI) | Time-Variant Relapse Error: Estimate (95% CI) |
|---|---|---|---|---|---|---|
| 3 | NSNE | 57.9% | 57.9% | 57.5% | 0.4% (-0.8 to 1.6%) | 0.4% (-0.8 to 1.6%) |
| | NSFE | 0.5% | 0.5% | 0.6% | -0.1% (-0.2 to 0.0%) | -0.1% (-0.2 to 0.0%) |
| | NSCE | 0.8% | 0.8% | 0.6% | **0.2% (0.1 to 0.3%)** | **0.2% (0.1 to 0.4%)** |
| | FSNE | 20.9% | 20.1% | 21.8% | -0.9% (-1.9 to 0.0%) | **-1.7% (-2.7 to -0.7%)** |
| | FSFE | 1.0% | 0.9% | 1.1% | -0.1% (-0.2 to 0.0%) | **-0.2% (-0.4 to -0.1%)** |
| | FSCE | 1.2% | 1.2% | 1.2% | 0.1% (-0.1 to 0.2%) | 0.1% (-0.1 to 0.3%) |
| | CSNE | 12.9% | 13.4% | 12.5% | 0.5% (-0.2 to 1.1%) | **1.0% (0.3 to 1.7%)** |
| | CSFE | 2.3% | 2.5% | 2.8% | **-0.6% (-0.8 to -0.4%)** | **-0.4% (-0.6 to -0.2%)** |
| | CSCE | 2.6% | 2.8% | 2.0% | **0.5% (0.3 to 0.7%)** | **0.7% (0.5 to 0.9%)** |
| 4 | NSNE | 56.1% | 56.1% | 56.0% | 0.1% (-1.2 to 1.5%) | 0.1% (-1.2 to 1.5%) |
| | NSFE | 0.9% | 0.9% | 0.8% | 0.1% (0.0 to 0.3%) | 0.1% (0.0 to 0.3%) |
| | NSCE | 1.1% | 1.0% | 0.6% | **0.5% (0.3 to 0.7%)** | **0.4% (0.3 to 0.6%)** |
| | FSNE | 20.6% | 22.4% | 22.7% | **-2.1% (-3.2 to -1.0%)** | -0.3% (-1.3 to 0.7%) |
| | FSFE | 1.5% | 1.5% | 1.4% | 0.0% (-0.2 to 0.2%) | 0.1% (-0.1 to 0.3%) |
| | FSCE | 1.5% | 1.4% | 1.2% | **0.4% (0.2 to 0.6%)** | 0.3% (0.0 to 0.5%) |
| | CSNE | 12.2% | 10.5% | 11.7% | 0.5% (-0.4 to 1.3%) | **-1.2% (-1.9 to -0.5%)** |
| | CSFE | 3.2% | 3.1% | 3.6% | **-0.5% (-0.7 to -0.2%)** | **-0.5% (-0.8 to -0.2%)** |
| | CSCE | 3.1% | 2.9% | 2.0% | **1.1% (0.9 to 1.3%)** | **0.9% (0.7 to 1.1%)** |

Static relapse rate STOP projections of smoking and e-cig use prevalence are compared against time-variant relapse rate STOP projections and empirical estimates from the Population Assessment of Tobacco and Health (PATH) survey. Static relapse STOP simulations have a constant relapse rate with respect to the number of months of abstinence, while time-variant STOP simulations have a relapse rate that decays exponentially with respect to the number of months of abstinence. Error is measured as the difference between the model-projected prevalence of each smoking and e-cig use state and the corresponding empirical prevalence in PATH.

NSNE = Never Smoker, Never E-Cig User
NSFE = Never Smoker, Former E-Cig User
NSCE = Never Smoker, Current E-Cig User
…
CSCE = Current Smoker, Current E-Cig User

**Bold** indicates that the p value of the error < 0.05

**Supplemental Figure S1.** Changes in transition rate estimates with respect to iteration of the weighted maximum likelihood optimization algorithm.

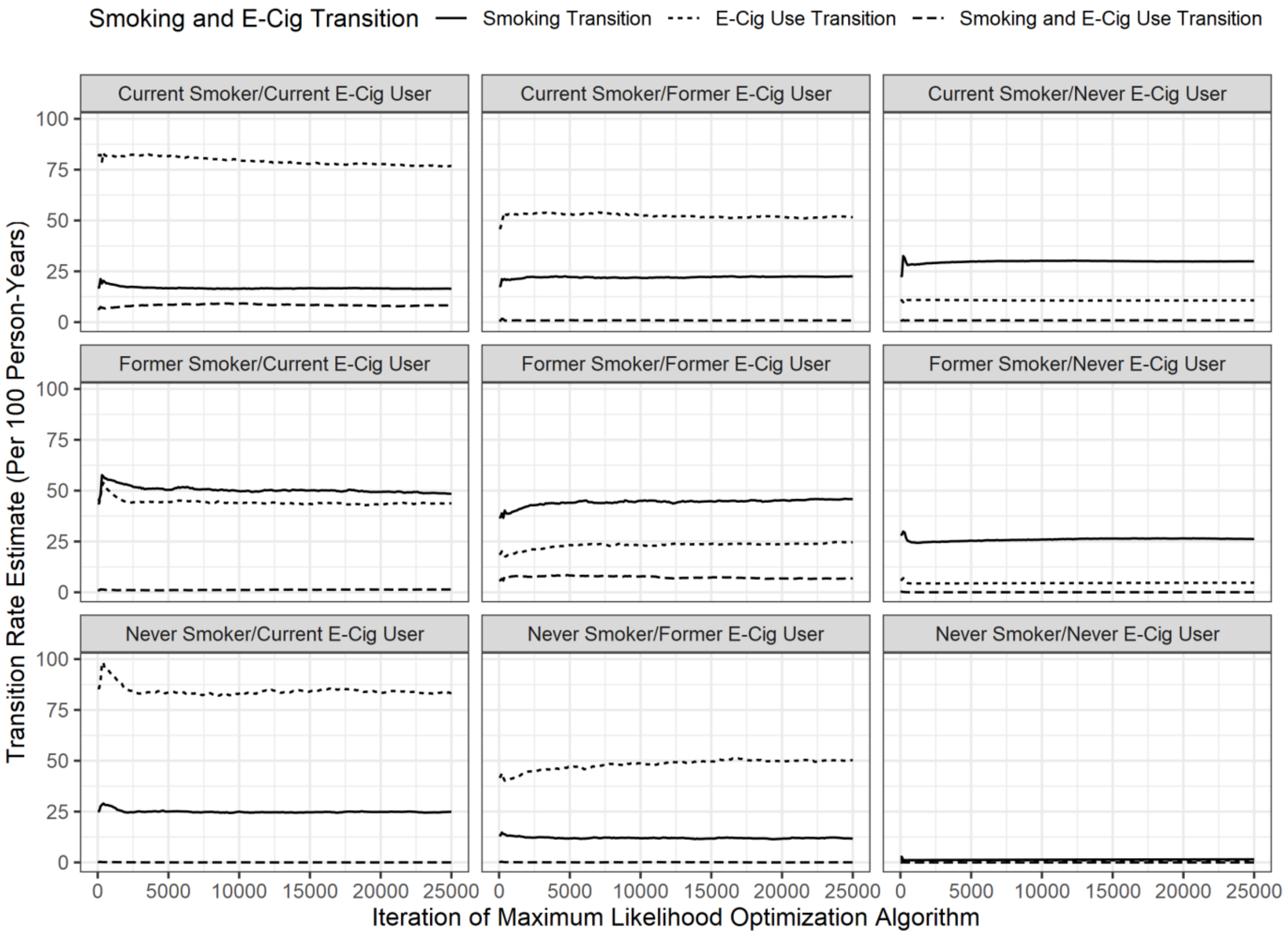

Transition rate estimates for each of the three possible transitions out of every smoking and e-cig use state are extracted from the continuous time Markov multi-state model every 100 iterations of the maximum likelihood optimization algorithm. The visualized rates are an average of age and sex-specific rates, according to their weighted prevalence in the Population Assessment of Tobacco and Health (PATH) data. Estimate convergence, as indicated by a relatively flat line, occurs before 25,000 iterations in all transitions.

**Supplemental Figure S2.** Monthly transitions categorized by type in the Simulation of Tobacco and Nicotine Outcomes and Policy (STOP) microsimulation model.

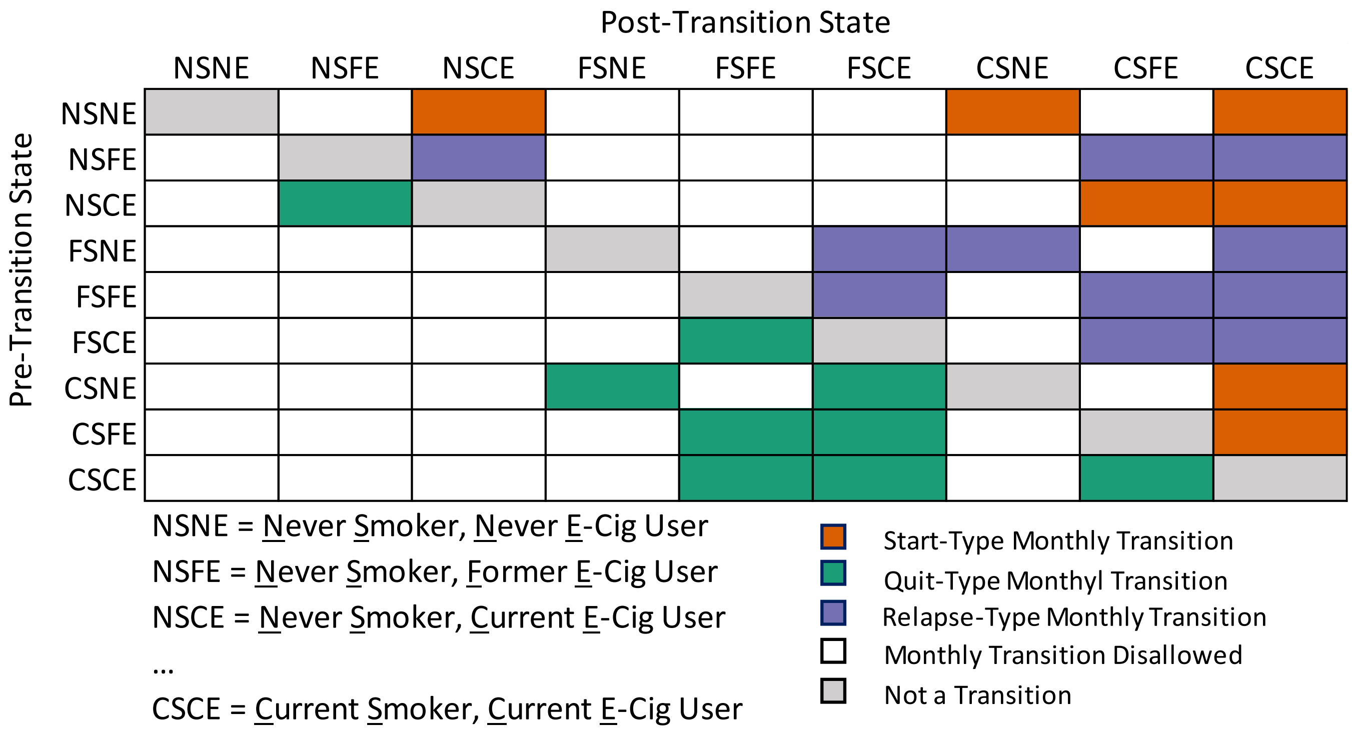

The STOP model allows no more than one transition to occur each month. Monthly transitions from the indicated pre-transition smoking and e-cig use state (row) to the indicated post-transition state (column) are categorized by type: start (red), quit (green), and relapse (purple). Start and quit transitions are stratified by age, sex, and current smoking and e-cig use state, but are constant with respect to time. Relapse transitions are stratified by age and current smoking and e-cig use state and have the option to decay exponentially with respect to duration of abstinence. Disallowed instantaneous transitions are in white.

**Supplemental Figure S3.** Validation of the Simulation of Tobacco and Nicotine Outcomes and Policy (STOP) microsimulation.

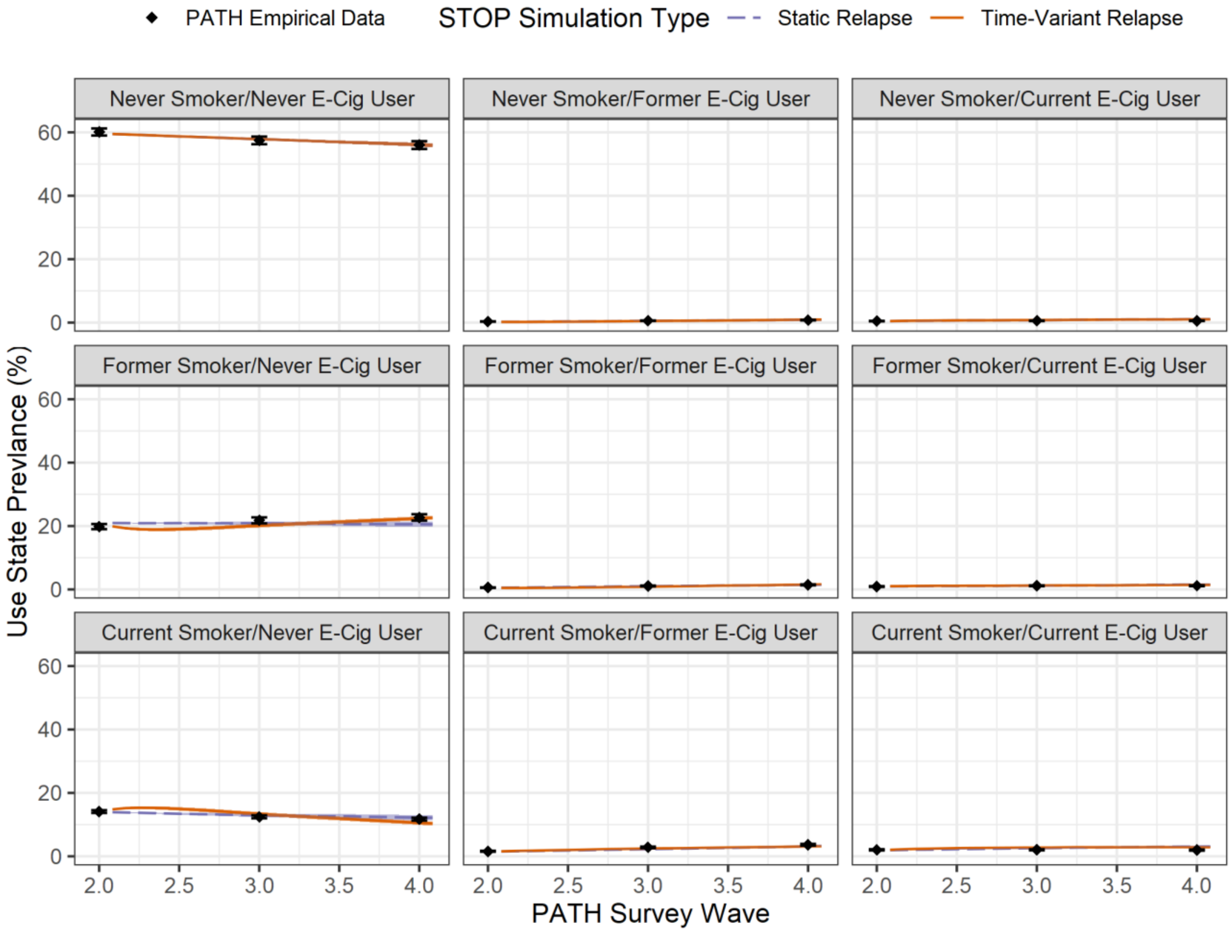

The STOP model is parametrized with Markov multi-state model-estimated smoking and e-cig transition frequencies, along with Population Assessment of Tobacco and Health (PATH) Wave 2 smoking and e-cig use prevalence. The static relapse simulation has constant relapse rates with respect to duration of abstinence, while the time-variant relapse simulation allows relapse rates among former smokers and former e-cig users to decay exponentially according to their durations of abstinence. STOP projected 12 and 24-month estimates of smoking and e-cig use prevalence are compared against PATH Waves 3 and 4. In some plots, the static relapse simulation line and the time-variant relapse simulation line are essentially superimposed. 95% confidence intervals are shown as shaded blue and orange segments for STOP static relapse and time-variant relapse simulations, respectively, and error bars for PATH empirical estimates. Confidence intervals are narrow enough to be nearly invisible in some sub-plots.
